# Rethinking supported self-management for Black people living with stroke: The relevance of social, cultural and historical racial context

**DOI:** 10.64898/2026.07.16.26357926

**Authors:** Jo White, Liz Livingstone, Mary Cramp, Emily Dodd, Tushna Vandrevala

## Abstract

**Objectives:** Despite their higher risk of stroke and known inequities in post-stroke outcomes, research amongst minoritised ethnic communities who have experienced stroke is scarce. This study aimed to explore the experiences of Black people in England and identify implications for supported self-management.

**Methods:** Between December 2023 and June 2024 qualitative interviews were conducted with 20 Black people living with stroke in England. Interviews were conducted in-person or online, depending on participants’ preferences. Data were analysed iteratively using reflexive thematic analysis.

**Results:** Three themes were developed: (1) “My world after stroke” capturing emotional and social identity-related impacts and adjustments; (2)“My support”: Family, Community and Peers, encapsulating different facets of support and the relational nature of self-management; and (3) “Supported self-management as a negotiated partnership with healthcare professionals”, highlighting the varied nature and outcomes of patient-professional interactions and their social embeddedness, including underlying power and historical racial dynamics.

**Conclusions:** Our study confirms the need for closer examination of how supported self-management can be provided to different populations. The post-stroke experiences of the Black people who participated in our study were inextricably shaped by their specific familial, social and cultural settings. Their adaptation to and management of the physical and emotional aspects of their condition was enacted across family systems, community and peer networks as well as healthcare services, with varying outcomes. In addition, their interactions and engagement with healthcare professionals were influenced by a broader historical context of discrimination and racism. Our study reveals the importance of delivering supported self-management to Black people living with stroke which responds to their specific social contexts and intersectional identities and which also strives to reduce power imbalances and address historical discrimination, thereby ensuring ‘cultural safety’ in service provision. Overlooking these aspects risks undermining self-management and underserving those who already face significant health inequities.

## Introduction

Stroke is a long-term condition disproportionately affecting Black communities in the UK, with incidence twice that observed in White populations (1) and onset occurring on average five years earlier (2). Risk factors, including diabetes, hypertension, and sickle cell disease (SCD), are more prevalent in Black communities (1, 3) with SCD increasing stroke risk in childhood and young adulthood (3). Ethnically minoritised communities experience higher rates of adverse outcomes post-stroke, including severe complications, pneumonia or mortality, and greater unmet recovery need (2). Specifically amongst Black African and Caribbean people, persistent care inequalities have been identified which are not fully explained by sociodemographic, cardio-vascular risk or stroke-related factors (4). Despite these inequities, the stroke experiences of Black communities are understudied.

Living with stroke presents multifaceted challenges, due to its sudden onset and physical, cognitive and psychological impacts (5). Many people report feelings of loss and struggling with their sense of self (6). Stroke also affects family members, particularly those who take on the role of carers. Wider social relationships are also impacted (7, 8).

Adjusting to life post-stroke involves adapting and developing coping strategies, with this process influencing subsequent quality of life (7). Stroke ‘self-management’ has been defined as “the actions and confidence… to manage the medical and emotional aspects of their condition, in order to maintain or create new life roles” (9). Supported self-management (SSM) is a central component of care for those living with long-term conditions (LTCs) such as stroke. SSM approaches focus on enhancing ‘self-efficacy’, derived from Bandura’s Social Cognitive Theory (10), conceptualised around a person’s belief in their ability to produce a given result from their actions (5). Four factors are understood to influence self-efficacy: previous ‘mastery’ (personal success), vicarious experience from observing others, verbal persuasion and psychological state (10). Concerns have been raised around the delivery of responsive SSM given a residual care model which prioritises expert-based knowledge (11, 12) while the longstanding emphasis on individual self-efficacy has been critiqued for not considering how social context affects a person’s self-management (5, 13). Self-management is increasingly recognised as not an individual endeavour but one embedded within social relationships (14), with family, peers, and wider networks playing an influential role (15–17). Indeed, on conceptualising self-efficacy, Bandura included the mechanism of ‘collective self-efficacy’ (10), namely shared beliefs and processes within group membership, including influence, perseverance and objectives towards behavioural outcomes. This collective process remains underexplored (16).

Uncertainties remain about how SSM should best be provided to different populations (18). Research with people of lower socio-economic status identified lower baseline self-efficacy, for example, attributable to vicarious experience of poorer outcomes, less control in everyday life, negative expectations, and resource barriers (13). These findings prompted recommendations for healthcare professionals (HCPs) working with such communities to “redefine and rediscover” SSM (13, p.960) which responds to social context, through taking a wider perspective of self-management objectives which provide hope and purpose, for example. It has, further, been argued that patient-HCP interactions should themselves be considered one element of the social context influencing self-management outcomes (11).

The facilitation of self-management has been embedded in UK stroke policy since the introduction of the National Stroke Strategy (19). SSM within stroke services aims to foster trusting relationships with patients and their families that enhance confidence and resilience (18). This responds to the recognition that tailored support is required (5), and patient-centred approaches adapted to individual circumstances can facilitate psycho-social adaptation (7).

Despite known inequities in post-stroke self-management outcomes amongst ethnically minoritised communities (20), few studies have examined their experiences. Evidence across a range of LTCs suggests, however, that people from such communities are less likely to participate in self-management interventions (21–23), and mistrust related to prior experiences of discrimination shapes their interactions with healthcare services (24). In the UK context, overall Black patients have reported lack of shared decision-making, poor communication and lower levels of satisfaction in services (23). Poor HCP awareness of and responsiveness to structural determinants of inequalities are also understood to weaken engagement with care (25). Such evidence has implications for the successful delivery of post-stroke SSM to diverse populations. Yet research on the factors influencing post-stroke experiences and outcomes is scarce.

This paper addresses current gaps in knowledge by presenting findings from a qualitative study which explored the experiences of Black people living with stroke (BPLS) in England. Through examining the role of social contexts we consider the implications for understanding self-efficacy, self-management and SSM. The study was part of an NIHR-funded project (INSERT DETAILS) that aimed to deepen understanding of the experiences and priorities of Black people living with stroke and co-design resources to support them.

## Methods

### Design

Our study employed semi-structured interviews to explore the experiences of BPLS. A qualitative design was appropriate given the study’s focus on subjective experiences and meanings following stroke within their social contexts (26).

### Participant Recruitment

Twenty-one participants were recruited as part of the wider co-design project (27). Purposive sampling was employed to ensure variation in age, gender, racial, community and socio-economic backgrounds, sickle cell status and clinical presentation.

**Table 1:** Demographic characteristics of participants with diagnosis of stroke.

|  |  |  |
| --- | --- | --- |
| Age | 30-39 | 2 |
|  | 40-49 | 2 |
|  | 50-59 | 7 |
|  | 60-69 | 6 |
|  | 70-79 | 2 |
| Age at first stroke | 0-9 | 1 |
|  | 30-39 | 3 |
|  | 40-49 | 5 |
|  | 50-59 | 7 |
|  | 60-69 | 3 |
| Gender | Female | 9 |
|  | Male | 10 |
| Self-Reported Ethnicity | Black British | 2 |
|  | Caribbean | 3 |
|  | Congolese | 2 |
|  | English/ Jamaican heritage | 1 |
|  | Ghanaian | 1 |
|  | Indian/Jamaican | 1 |
|  | Nigerian | 7 |
|  | Ugandan | 1 |
|  | Zimbabwean | 1 |
| Education | Secondary School | 4 |
|  | NVQs | 1 |
|  | Degree | 9 |
|  | Post-Graduate | 3 |
|  | Not given | 2 |
| Work Status | In work | 5 |
|  | Unemployed | 1 |
|  | Retired | 4 |
|  | Not working due to acquired disability | 9 |
| Living Arrangements | Lives alone | 6 |
|  | Lives with family | 10 |
|  | Lives with family, has formal carers | 3 |

‘Living with stroke’ was defined both as those who had experienced a stroke and carers. Participant recruitment was facilitated by community-based and national organisations providing health support to Black communities and professional contacts within the research team. This approach enabled us to successfully recruit a diverse mix of BPLS who may be less visible within formal healthcare pathways, although introduced a partial bias towards those already connected to either community or national organisations. However, personal contacts through our PPI group enabled us to recruit several participants who were more socially isolated. Ethical approval was granted by (REFERENCE TO BE INSERTED). Initial contact with participants was made by researchers via email or telephone, through social media channels employed by national organisations, and face-to-face at community groups. Recruitment materials were produced in an aphasia-friendly format to enable inclusion of those living with aphasia. All participants received study information and provided informed consent prior to participation. All participants were compensated for their time with an e-voucher of £15.

### Data Collection

A semi-structured interview guide was developed with input from PPI group members and refined through two pilot interviews (SEE ADDITIONAL FILE 1). The questions were designed to encourage participants to reflect openly on their post-stroke experiences from initial stroke onset to hospital care and rehabilitation and return home, including participants’ early reactions, how they adapted, their social context and relationships, the sources of support received, their interactions with HCPs, and their perspectives about living well with stroke. We deliberately chose not to use the term ‘self-management’ directly with participants, given this term is not part of everyday language amongst those living with stroke but is more associated with services.

Interviews were conducted between December 2023 and June 2024 and lasted between 40 and 120 minutes. They were carried out face-to-face in a semi-public place, in participants’ homes, online via Microsoft Teams, or by telephone, depending on each person’s stated preference. All interviews were conducted in English. While project funds were available for interpreter support in other languages, but this was not necessary.

Five members of the project team were involved in data collection: three university researchers and two community researchers. These were a White British female postdoctoral Research Fellow with experience in qualitative research methodologies and stroke self-management, a Chinese female postdoctoral Research Fellow experienced in qualitative research methodologies and service user engagement, and a White British female Senior Research Fellow with expertise in qualitative research and public involvement. All academic researchers underwent a reflexivity exercise early in study planning to identify and work to address any biases and assumptions informing their activities. The two community researchers (CRs) were female and from Black communities; one was a carer of a family member who had experienced stroke. They received training in qualitative methods, ethics, confidentiality, reflexivity, and safeguarding. One CR accompanied a university researcher to three in-person interviews, contributing to the interviewing process, and one CR led an in-person interview individually. All other interviews were conducted individually by academic researchers, which may have affected rapport. The interviews were recorded, transcribed and anonymised prior to analysis.

While our support of both in-person and online interviewing promoted inclusion, our experiences of online research confirmed the risk of fraudulent participation. This phenomenon is exacerbated by the use of social media channels for study recruitment and is now so widespread that risk management strategies and proactive preventive measures have been recommended (28). Eight respondents recruited through social media channels exhibited unusual behaviour during online interviews, such as declining to turn on their cameras, offering similar (yet atypical) descriptions of stroke symptoms, offering one-word answers to open questions, or using generic language throughout their responses. Invalid telephone numbers and postal addresses were frequently provided following our request for additional contact details, as required by university finance departments for e-voucher disbursement auditing. In one case a consent form from a different, unrelated online study was submitted to researchers by mistake. The transcripts from these ‘suspect’ interviews were excluded from our final sample of 20 interviews following team discussion and consensus that these constituted fraudulent participants.

### Data Analysis

Data were analysed using reflexive thematic analysis, an approach suitable for exploring subjective lived experience trajectories, while acknowledging the role of researchers in knowledge production (29). Following data familiarisation, codes were generated independently by two researchers (XX and XX), focusing on how participants described managing life after stroke, including social relationships and context and relevance to self-efficacy and self-management. Codes were iteratively refined through review and discussion with the third researcher (XX). This enhanced the methodological and interpretive rigour of the analysis and enabled consensus in interpretation, although a larger, more diverse team might have identified additional patterns within the data. The finalised list of codes was then assembled into candidate themes which were developed into final over-arching themes and sub-themes through continued discussion amongst the three researchers. The study was conducted in accordance with a consolidated criteria for reporting qualitative research (COREQ), enhancing transparency and transferability and providing an audit trail for all activities (SEE ADDITIONAL File 2).

## Results

Three overarching themes were developed, each with a set of sub-themes: (1) “My world after stroke” capturing emotional and social identity-related adjustments; (2)“My support”: Family, Community and Peers, encapsulating facets of support and the relational nature of self-management; and (3) “SSM as a negotiated partnership with healthcare professionals”, highlighting the varied nature of patient-professional interactions, their social embeddedness and the power and historical racial dynamics underlying them. Together, these themes illustrate how self-management is not just experienced as an individual endeavour but is inextricably shaped by social context.

### My world after stroke

Participants described their initial reactions and the process of adapting to their situation.

#### Shock and grief for my old self

Many detailed the vivid emotional impacts of stroke; their “life turned upside down” as one participant expressed it. As stroke was understood as predominantly affecting older people the shock experienced by younger participants was particularly profound. Feelings of loss were recalled during the struggle to reconcile pre- and post-stroke identities. Coming to terms with the ‘new normal’ was described as a long, arduous journey.

> *I think it’s a hard position to accept that you’re no longer who you were (P12, female in their 40s)*

For younger BPLS - particularly those balancing employment and parental responsibilities - the new situation was intensely disruptive.

> *We are young people; you’ve got a lot going on. Kids, work, family (P20, female in their 40s, wife of P19)*

Participants highlighted not only new physical limitations but distress, guilt and a sense of inadequacy at their inability to fulfil previous family roles.

> *I was the main breadwinner… I had children (P1, female in their 60s*)
>
> *My children when they say, ‘mommy, when are you going to get better?’ When they ask me that, I will start crying… I’m a woman, I used to cook for my family…’ (P2, female in their 50s).*
>
> *Emotionally, that is difficult, especially with having my young boys, because you feel guilty about being limited on the things that you can do with them… (P17, female in their 30s)*
>
> *These early experiences of grief and disrupted identity often undermined confidence and capacity, fundamental components of self-management.*

#### Learning to live with my new reality

As participants moved beyond initial shock and sorrow, they described a - sometimes uneven - process of adjustment. This included finding ways of coping emotionally and reclaiming some control. For many, self-management involved redefining independence in the light of their changed abilities. Several referred to the importance of maintaining continuity with aspects of their pre-stroke roles and identity to *“claim as much of their life before the stroke as possible”* (P1, female in their 60s). This meant finding alternative ways to engage in valued activities, rather than abandoning them.

> *Don’t give up the things you love completely… There’s a different way of doing what you used to love* (P20, female in their 40s, wife of P19).

Younger participants described how both physical and emotional adjustment were frequently impeded by health services (e.g., stroke wards and rehabilitation settings) designed primarily for older adults. Rehabilitation targets were often set well below their abilities, for example, which meant that they found themselves not meeting acceptance criteria for services and support, despite their important personal goals, such as returning to work.

> *It’s so minimal that the rehab when I looked into our local services… It was shocking … it wasn’t specific enough for what the needs* were (P19, male in their 40s)

Concerns were expressed around formulaic, ‘tick box’ care and its impacts, not only on self-management amongst younger BPLS, but for all patients.

> *I think everybody was lumped together. I didn’t notice any difference when we were in hospital, in rehabilitation or anything… seeing other people around was just awful.. to see what support they were getting and not getting, and the effect of that on them whilst they were there was just something else (P20, female in their 40s, wife of P19)*The ability to adapt and self-manage was also influenced by contextual factors such as the availability of transport and family members who could offer support.
>
> *I think they had some service where you had to go there, like was it like once a week or something? … But I didn’t take that up though. Trying to get to that location was tough* (P19, male in their 40s)

Participants’ accounts illustrated how adaptation involved adjusting both physically and emotionally and redefining engagement in meaningful activities. However, this process was frequently constrained by external as well as individual factors, highlighting the significance of systemic and structural barriers.

“My support”: Family, community and Peers

For most participants, adapting to life post-stroke was described as a shared, relational process. Families and social networks - friends, peers, community - provided critical practical, emotional and advocacy support.

#### Families go through this together

Many participants described feeling “held” by those around them. Relatives not only provided support at home and in daily life but often helped them navigate complex health systems, served as intermediaries with HCPs, and worked to ensure their needs were recognised and supported. Partners, parents, and children often adjusted their own lives to accommodate new caring responsibilities. One participant had his stroke six days after his youngest child was born. His wife described balancing parenting a newborn with advocating for her husband.

> *I spent the next three months at the hospital with the baby … because that was all I was worried about… his care*… *My mum took early retirement and came ’cause she saw how, you know, how bad things were* (P20, female in their 40s, wife of P19)

The new situation had emotional repercussions on all family members.

> *I remember the first time my son saw [his] Dad again… He looked very different by then. He ran in… he didn’t want to go further … And he was only two, but he knew something’s up. “I’m not going near that man” … I wasn’t prepared for that, how the children would deal with it*

Supporting their children’s adjustment involved ‘normalising’ his father’s rehabilitation and reframing it as a shared family activity, with positive outcomes.

> *When trying to do, like, some exercise at home and stuff like that, I include the kids… We can try and make it fun… She [the baby] was [redacted] days old when it happened. They learnt to walk again together* (P20, female in their 40s, wife of P19)

In other cases, the shift in intra-family responsibilities and its impact left longstanding feelings of regret.

> *I feel like my stroke took my daughter’s childhood away, she grew up too quickly and too fast. Because she lost things we used to do* (P3, female in their 60s)

Despite the vital role played by families, several participants highlighted the lack of recognition of this, and of the informational and wider support needs of family members, within stroke services.

> *I don’t actually remember the NHS doctors coming in and asking the [my uncle’s] wife, ’how are you doing?‘* (P11, nephew of male in their 70s)
>
> *At what stage were you listening to us? What stage was communication happening? None. Listen to the person, listen to their families. What support network do they have?… You know, the families are impacted … involve them in their in their care plan* (P20, female in their 40s, wife of P19)

For those who were recent migrants, limited family members in the UK added a further layer of complexity, with extended family or community networks compelled to take on caring responsibilities; a significant, unexpected disruption.

Family members were therefore, central to the enactment of self-management, but their role was rarely recognised within SSM. Conversely, some participants reported having no or limited family support, with family unavailable, unable or unwilling to play an active caring/supportive role. This left them isolated.

> *I managed to go [to stroke support group] about three times … because I didn’t have nobody to take me… my sister has to work… (*P14, male in their 50s)

#### Communities support and silence

Community, including faith and social groups, was central to many participants’ lives, both before and following stroke, and in many cases provided invaluable support.

> *INT So you had a good community to support you?*
>
> *R: Yes. That was why I survived this thing* (P2, female in their 50s)
>
> *We’ve just been so blessed of the support we’ve had around us from our church to our families* (P20, female in their 40s, wife of P19)

However, others described a reluctance to talk about stroke and its wide-ranging impacts, including effects on emotional and mental health, within their communities. Prevailing secrecy and shame inhibited them from seeking support.

> *I know that within our community… we don’t talk to people about our feelings and what we’re going through, and we don’t access support, necessarily, unless it’s in crisis …* (P17, female in their 30s).

In addition, several participants described how stigma and misconceptions around stroke inhibited disclosure.

> *My family, my parents especially, still haven’t gotten over the trauma of me having a stroke. They haven’t even told people back home… It’s that bad, it’s like it’s a slight stigma. And people think that stroke is for the older people* (P3, female in their 60s)

In some cases, cultural explanations about stroke within their community led to poor advice and self-blame.

> *Our community, or our friend they say ‘you got a stroke because you are thinking too much’. I said ‘okay’… I believed people when they said that* (P8, male in their 60s)
>
> *Because for Africans, they think when you got a stroke it’s because you are thinking too much. You got a thinking problem … the advice they give to you, they said, ‘my friend, try to smile a lot’… It’s like, stroke is from stress; they got that in their head. It’s very hard to change* (P6, male in their 60s)

Some participants expressed reluctance to socialise in their communities due to negative expectations.

> *The way I look, you know, my community, I don’t like going … some people are going to start talking, talking. I don’t like that. I started to prefer to stay home myself … I can go to my own family … it’s okay. But invite me to event like a party for everybody; I’m not going.* (P8, male in their 60s)

In this case the participant’s self-consciousness about his physical changes and anticipated reactions inhibited his behaviour. In contrast, several participants highlighted the power of initiating conversations about stroke, to overcome stigma and obtain necessary support from their communities, while also acknowledging barriers to communication.

> *If you can have open and honest dialogue and tell people what was going on, then they can help you in ways that you need… I know that sometimes that’s scary, but… people can’t help you if they don’t know anything’s wrong…* (P17, female in their 30s).

These findings reveal how community contexts both enabled and constrained self-management, shaping if and how individuals shared experiences, socialised, and accessed support.

#### Support from people like me

Peers can provide positive vicarious experiences which enhance the development of self-efficacy. Participants described how highly they valued the support and inspiration that they had drawn from meeting others who were also living with stroke and shared personal insights.

> *In the group people tell you different things about their experiences of what’s happened to them. And then sometimes you listen to them and then you can pick some things as well* (P4, female in their 60s)

Participants, further, described how seeing ‘people like me’ in these groups was particularly helpful in making them feel comfortable enough to ‘open up’. Conversely, they found it harder to engage with groups that did not include others of similar ethnicity and/or age.

> *My husband, has taken me to the After Stroke groups which is good … actually, there’s more black and brown people in there now, but there wasn’t to start off with, (P7, female in their 50s)*.
>
> *It’s alright going to the other groups, it’s just that they’re all quite a bit older, and like … it’s a different experience for me - especially having young children as well - than it might be for someone who’s older … might be retired. They don’t need to think about going back to work* (P17, female in their 30s)

Our findings highlight the importance of relatable role models in terms of age - and associated life stage - as well as ethnicity who offered collective sharing and enabled the development of self-efficacy through vicarious experiences.

Supported Self-management as a negotiated partnership with healthcare professionals

SSM requires HCPs to cultivate collaborative relationships with patients and their families and foster knowledge and skills to manage live post-stroke. Participants’ accounts revealed wide variation in their interactions with HCPs, yet these experiences played a fundamental role in shaping both their confidence and capacity.

#### Experiences with HCPs enable or inhibit self-management

Positive interactions were characterised by open communication, a recognition of personal experiences and concerns, and an effort to reduce power imbalances. One participant emphasised how her GP had given her time and space to talk about her situation holistically and discuss potential solutions.

> *She is just somebody that I could speak to and explore my health issues and how my health issues are affecting the rest of my life, and what I could do about it…*
>
> *I walked out of there feeling a lot more in control; a lot more empowered* (P1, female in their 60s)

When HCPs created a safe space for dialogue, participants reported feeling more confident to ask questions, share and discuss concerns, and “work” together in partnership.

> *They [rehab therapists] acted as if we are friends, and you can trust us… And so that makes you feel so easy… easy and free to ask them anything that is bothering you… Because if you’re working with someone and they trust you, they are able to tell you everything* (P4, female in their 60s).

Positive interactions validated participants’ experiences and responses.

> *They’re really good at telling you what… normalising a lot of it… and also telling you, like, hints and tips to do stuff, and talking about things like boom and bust and having to pace myself [to prevent fatigue]. The neurovascular nurse practitioner, is really helpful, cos she’s very validating… “Yeah, it’s okay that you feel that way”* (P17, female in their 30s).

Here, a personalised approach, combined with the provision of practical strategies for managing everyday challenges, is an example of ‘verbal persuasion’, crucial to self-efficacy.

The various experiences reported illustrate how healthcare interactions can foster trust and shared decision-making through HCPs creating space for dialogue, acknowledging patients’ broader life contexts and experiences, and working to reduce traditional power imbalances. Contrastingly the negative encounters reported were marked by an explicit power imbalance and lack of two-way exchange, with patients not feeling heard or seen as individuals.

> *I was not listened [to]. Because I find that some medical professionals, most of them… they put themselves on this high pedestal* (P3, female in their 60s)

Across multiple accounts, service provision was described that did not include the sharing of key information and assumed prior knowledge about stroke causation and prevention and the capacity to navigate complex information and health systems. This did little to support patients and families who were already managing new, uncertain roles and emotional strain.

> *One thing they should understand is we all come from different backgrounds and different cultures… And we have to learn from each other* (P4, female in their 60s)
>
> *A lot more information and more clarity [was needed] … I’m not sure if it was assumed that we knew? Most of the time I had a feeling… that practitioners or professionals just assume that’s information that we would have known (P11, nephew of male in their 70s)*

Several participants who had been unaware of the links between co-morbidities and stroke and had neglected to take prescribed medications were subsequently told ‘this is why you had your stroke’ by professionals. Consequently, they experienced feelings of guilt and self-blame, which made adapting to and accepting their situation even harder. Where interactions were perceived as hierarchical, dismissive, lacking cultural understanding or compassion, participants described feeling unheard and disempowered.

#### Historical backdrop shaping interactions

Some participants described how exchanges with HCPs intersected with historical experiences of racial discrimination and could not be separated from a broader social and historical context, which shaped both expectations of care and perceptions of how they were treated. For example, some questioned whether delays in treatment and access to specific medication were influenced by their race and not individual need.

> *I think if I was white they would’ve sent me for an MRI scan… If I was given all the resources, all the tests, they would’ve stopped my stroke … We are people, we are human beings. You cannot put people into boxes, thinking that this medication works well with Black people. No, treat everybody as an individual because we’re all unique* (P3, female in their 60s)

Some participants explained how previous poor personal experiences, including racial discrimination, contributed to a lack of trust and engagement in services.

> *I think that [previous bad experiences] does cause a lot of mistrust between us [Black] patients and healthcare professionals. Obviously if there is that mistrust… unfortunately it might be that they didn’t come to hospital in time because they know they’re going to get treated really badly… that you do end up having really adverse outcomes* (P10, female in their 30s).

The underlying (and historical) context of racism formed a critical, yet often unacknowledged backdrop to participants’ interactions with HCPs. For example, one participant described her vigilance around professional responses, linking these to racial bias.

> *I find that as a Black person you could just tell somebody who is warm towards you or somebody who isn’t… (P3, female in their 60s)*

Another reported moderating her behaviour with HCPs, to avoid being labelled an ‘angry Black woman’ – a self-censorship to avoid conforming to negative racialised stereotypes.

The racialised context of participants’ lives informed both how they monitored and interpreted interactions and social dynamics and how they responded to HCPs as they navigated health services. The heightened sensitivity reported can be considered a form of emotional labour within health encounters and is a crucial element of the wider social and historical context affecting BPLS.

Some participants explicitly emphasised a requirement for HCPs to (re-)build trust and reassurance based on an understanding of this wider context affecting Black communities and their historical relationships with services and institutions.  

> *… because the Black community in particular, they need reassuring giving the historical backdrop of everything that has happened. So professionals need to be aware of that* (P11, Nephew of male in their 70s).These findings emphasise not only the need for trust-building approaches from HCPs which facilitate open communication, but illustrate the need for service provision grounded in and responsive to understanding of wider historical factors and dynamics.

## Discussion

Despite known disparities in stroke risk and inequities in post-stroke self-management outcomes amongst ethnically minoritised communities (20), research exploring their experiences is scarce. This study with BPLS in England explicates the relevance of individual, social, relational and wider institutional and structural factors to stroke self-management and SSM. Participants’ adaptation to and management of the physical and emotional aspects of their condition was a relational process enacted across family systems, community and peer networks and healthcare services, with varying outcomes. Our findings align with and elaborate existing critiques of individualised models of self-efficacy which have emphasised how social context and inter-personal relationships, including the role of HCPs, all affect self-management (11). They also build on previous research with ethnically minoritised communities which challenged traditional theoretical framing around self-efficacy and revealed the power of familial and wider collectives in achieving behavioural change (15).

Consistent with existing literature, our participants described profound experiences of loss, disruption and identity change post-stroke (6), and a ‘ripple effect’ on family members. Family not only provided physical and emotional support but played a crucial role in advocating for care, a role which was not always accommodated by services, highlighting the importance of family carers being systematically included in decision-making, as part of collaborative SSM (18). In instances where family were unavailable, unable or unwilling to assist, impacts of stroke were more acutely experienced in terms of social isolation and capacity to engage with support.

For the BPLS in our study, meeting and relating to others who had experienced stroke and hearing about their self-management strategies and achievements was highly motivating. A key issue regarding peer support was being able to engage with others of a similar (often younger) age and racial background, underscoring the crucial intersectional aspects of post-stroke identity. For those who experienced stroke at working age it was important to meet peers who shared similar goals around returning to employment, for example. Maximising peer support has been recommended for post-stroke self-management interventions to enable vicarious learning and increase motivation (16). Less attention has been paid to the need of those from ethnically minoritised communities for peers who are relatable beyond shared experience of any given condition, due to wider common identities. Our findings on the value of peer group support also highlight the relevance of ‘collective efficacy’, namely shared understanding and group influence and perseverance towards behavioural outcomes (14, 16). While this concept has been less explored in theoretical examinations of self-management to date, it merits greater investigation (14). Further research which explores what constitutes ‘shared understanding’, vicarious learning and group influence, focused on ethnicity, age, gender, employment, socio-economic status and other factors relevant to BPLS and other minoritised communities is recommended.

Our study highlighted the varied role and impacts of communities founded on shared cultural affinity and faith. While some participants characterised such community networks as vital sources of support, others highlighted stigma, silence and misconceptions which impacted social isolation and mental health. Community dynamics shaped disclosure and support seeking, impacting self-efficacy and self-management. HCP understanding of the effects of community context(s) on those living with stroke is critical to the provision of responsive SSM.

Our findings also offer new insight into how HCP-patient exchanges and relationships are a key aspect of the broader social context influencing the experiences of BPLS. Interactions with HCPs constitute a mechanism through which self-management is either enabled or inhibited. Participants’ accounts highlighted how SSM can be understand as a process negotiated through the relationships established by HCPs. Where there was open communication, efforts to reduce power imbalances and validation of their experiences, participants felt more trusting, confident and empowered in discussions about managing their condition. Contrastingly, where a hierarchical and unhearing approach was experienced, interactions were disruptive, posing an obstacle to service engagement and confidence-building, impeding self-management. This demonstrates how SSM and its effectiveness is inextricably tied to relational dynamics.

A further contribution of our study is its explication of how BPLS-HCP interactions are shaped by a broader social and historical context of discrimination and racism. Our findings confirm how this context not only creates mistrust and negatively impacts expectations (13), but also has a recursive effect, impeding authentic communication. Hence participants described heightened vigilance and, in some cases, modifying their responses to HCPs to avoid negative reactions based on racial stereotyping. Such dynamics place an ‘additional burden’ on patients. While conceptualisations of self-management and SSM have highlighted the need for personalised, collaborative approaches in response to familial and social contexts, these findings highlight the need to also consider wider historical racial contexts. Previous research has posited that psychosocial and structural determinants may be contributing to the major and persistent ethnic inequalities in stroke care and outcomes identified amongst Black African and Black Caribbean people, given current disparities are not fully explained by sociodemographic, cardio-vascular risk or stroke-related factors (4). Our research confirms how different social contexts can facilitate or hinder psycho-social adaptation (7) and, further, suggests that psychosocial and structural aspects are inextricably interlinked, given the wider historical context within which service delivery is situated and experienced.

Our study highlights the importance of HCPs creating person-centred approaches which involve closely listening to patients’ experiences, developing shared learning and meaning and ensuring interactions offer respect and dignity, while working to remove traditional power dynamics. It also confirms how HCP understanding of the historical inequities affecting minoritised communities and service provider awareness of broader structural determinants influencing patient (dis-)engagement is critical to developing mitigatory action to address health inequalities (25).

Our findings support calls for closer examination of how SSM should best be provided to different populations, given prevailing uncertainties (18). Prior research with those of lower socio-economic status has highlighted lower baseline self-efficacy, linked to vicarious experience of poorer outcomes, less sense of control in their everyday lives and negative expectations (13). Our research reveals how recognising the significance of specific social and historical contexts and taking a social-informed view of the aspirations of underserved communities is required. In the case of BPLS there is a need for informed care which is personally and culturally responsive and a trust-building approach to SSM, grounded in an understanding of wider historical and structural inequities faced by Black communities.

In terms of implications for practice and policy, our findings point to the value of introducing ‘cultural safety’ concepts to SSM. Cultural safety (CS) is founded on principles of tackling historical impacts of marginalisation, discrimination and institutional racism and interest in this approach is growing, both in the UK and internationally (30). CS is a recommended approach for delivering care to ethnically minoritised groups through the creation of person-centred experiences, removal of traditional hierarchies and improved institutional facilitation and structural reflexivity (30). Modifying policy and practice by institutionalising training and mentoring for staff which incorporates CS principles, can make an important contribution to the conceptualisation and delivery of SSM. Conversely, models of SSM which overlook social context and relational dynamics, including wider historical power and racial structures, risk underserving those who already face significant health inequities.

## Supporting information

Supplemental File 1 Interview Guide

Supplemental File 2 COREQ checklist

## Data Availability

The data analysed for the current study are available from the corresponding author on reasonable request.

## Notes

### Competing Interest Statement

The authors have declared no competing interest.

### Author Declarations

This study received ethical approval from the University of the West of England (UWE Bristol) Research Ethics Committee (Ref CHSS.23.10.044).

### Funding

- National Institute of Health and Social Care Research (NIHR): NIHR204975

