## Supplemental File 1 Interview Guide for "Rethinking supported self-management for Black people living with stroke: The relevance of social, cultural and historical racial context"

### **ADDITIONAL FILE 1**

### ISSMAS Project

### Interview guide for Black People Living with Stroke

*Put participant at ease.*

***Confirm capacity***

- ***Check they have read and understood the information sheet***
- ***Ask them what they understand the study to be about***
- ***Confirm what their interest in participating.***

*Ask if they have any questions about the study*

*Check that they have signed the consent form.*

*Confirm the interview will be recorded, but identifying details removed from transcripts.*

*Remind them the interview may take up to an hour, but they can request for it to be paused if they need a break and can end it at any time.*

Would you mind telling me about your current living situation?

*Prompts: Who do you live with? Do you look after your day-to-day needs or do you have someone/others helping you? Confirm role of family/friends/formal carer.*

***Stroke and early self-management***

When did your first stroke happen?

Have you had any further strokes, and if so, when did these happen?

Did you realise at the time that you were having a stroke?

(If yes – how did they know, where did that knowledge come from).

Thinking back to your first stroke, did you have any pre-existing conditions or early physical symptoms which you now understand were to do with your stroke?

If yes: Please elaborate

***Hospital stay and returning home***

How long did you spend in hospital following your stroke?

Can you remember any	information and advice you were given before going home?

*Prompts: Lifestyle changes to support your health, e.g. diet, exercise, medical support (ask for details/examples); Sources of support (details/examples)*

How was this information given to you/your carer?

What opportunities were you given to ask questions before being discharged?

*Prompts: Who did you speak to? (which HCPs); Did you feel comfortable asking questions?*

*What might have helped you ask (more) questions and discuss your concerns?*

Did you or your carer have any concerns about how to manage back at home after leaving hospital?

Was there any information, advice or support you or your carer would have liked to have received then which you felt was missing?

If yes: What would you have liked?

**CHECK RESPONDENT IS NOT TIRED OR DISTRESSED AND IS FINE WITH CONTINUING**

***Follow up after stroke***

Have you had any reviews or regular check-ups since your initial stroke diagnosis?

*Prompts: 6 month review; annual review with GP; appointments with other health and care providers*

Are you able to talk about the things that are important to you in these reviews?

If not – what would you have liked to be able to discuss?

What do you think might help you be able to talk about things that are important to you?

***Reflections on health and care service provision/communication***

Do you think that anything about you (e.g. your gender or race, ethnicity or cultural background or any other aspect) has affected how you have been treated within health and care services since your stroke?

If yes: Could you describe how?

***Emotional/ psychological impacts and needs***

Do you think that your emotional wellbeing or mental health have been affected by having a stroke? If so, in what ways?

*Prompts: Shock, fear, anxiety or depression at different stages; change in sense of self; change in relationships.*

Have you received any kind of advice or support to help you with your emotional wellbeing and mental health?

If yes: Where did you get this from? How has it helped you?

If no: What would you find helpful?

**CHECK RESPONDENT IS NOT TIRED OR DISTRESSED AND IS FINE WITH CONTINUING**

***Living with stroke – your self-management now***

Does your stroke affect your day-to-day life now? In what ways?

What are your main concerns at the moment?

Would you say that you now understand what caused your stroke and how to reduce your risk of further stroke?

*Prompts: Information sources/providers which have aided understanding; any gaps in understanding.*

Do you feel that you/your carer have made any changes to your life to reduce your risk of further stroke?

If yes: What are these?

What/who gave you the idea to make these changes?
What motivated you to make these changes?

What/who helped you overcome any difficulties in making these changes?

Have you or your carer made any other changes to help you manage things?

If yes: Can you explain these, who made them, and the difference they have made?

Are there any sources of information you would you say you most trust about stroke?

*Prompts: Online medical/online organisation (trusted sites), health/medical community/ community organisation/information-sharing in-person.*

What is it that makes you trust these particular sources?

Given what you know now, what advice would you give other people who have experienced stroke about how to manage things following stroke?

**CHECK RESPONDENT IS NOT TIRED OR DISTRESSED AND IS FINE WITH CONTINUING**

***Views on Resources for Black People Living with Stroke***

In our project we are developing information and guidance to help people from the black community living with stroke.

Is there anything you think people in your community need to know about stroke which we should include?

How do you think this information and guidance should be provided?

*Prompts: What kinds of media/format? Use of text/visual media? Short videos? How should these present information? Special leaflets? How should information be presented?*

How do you think we should make any new information and guidance (e.g. leaflets/videos) available to people in your community?

*Prompts: best ways of dissemination*

***Views on Resources/Intervention for Health and care professionals***

In our project we are also planning to develop information and guidance for health and care professionals (HCPs) to help improve the support they provide to black people who experience stroke.

What do you think HCPs need to know about your experiences/the experiences of black communities in order to provide a better service to you?

How do you think health and social care professionals (eg. hospital staff, your GP, or pharmacist) could provide better information and support?

*Probe: Experience/reasoning here – kinds of information/style of communication/support desired*

***Confirming background details***

***Check here for any contextual details not provided informally at the recruitment stage or in the course of the interview***

**I’d like to check some background details about you. We ask these questions to check that we are representing a broad range of people in our research – but if you feel uncomfortable with a question, you don’t have to answer**

Where are you currently living (which town/city)?

What is your current job/work situation?

Has this changed since having a stroke?

If yes: Please share details

Can you tell me a little about your education? (What age did you leave school and what did you do after that?)

Do you currently have any medical conditions you are aware of?

*Prompts: Sickle cell, Type 2 diabetes, high blood pressure, high cholesterol*

Please can you tell me your age?

What gender do you identify with?

How would you describe your racial and/or ethnic background?

Thank you. We have nearly finished the interview now.

Is there anything else that we haven’t talked about that you think is important to tell me?

Thank you so much for your time.

*End interview*

*Outline possibility of follow-up interview, next steps of project, and opportunities for further involvement*

*Remind respondent that they can request to withdraw within the next two weeks and their data will be withdrawn from the project.*
